# Public Health and Social Measures (PHSM) interventions to control COVID-19 An Overview of Systematic Reviews

**DOI:** 10.1101/2023.11.21.23298387

**Authors:** Racha Fadlallah, Fadi El-Jardali, Nour Kalach, Lama Bou Karroum, Reem Hoteit, Andrew Aoun, Lara Al-Hakim, Francisca Verdugo-Paiva, Gabriel Rada, Atle Fretheim, Simon Lewin, Ramona Ludolph, Elie A. Akl

## Abstract

Response to the COVID-19 pandemic included a wide range of Public Health and Social Measures (PHSM). PHSM refer to a broad array of nonpharmaceutical interventions implemented by individuals, communities and governments to reduce the risk and scale of transmission of epidemic- and pandemic-prone infectious diseases. In order to inform decisions by the public, health workforce and policy-makers, there is a need to synthesize the large volume of published work on COVID-19. This study protocol describes the methodology for an overview of reviews focusing on the effectiveness and/or unintended health and socio-economic consequences of PHSM implemented during the COVID-19 pandemic. Findings can shape policy and research related to PHSM moving forward.

## Background

Coronavirus disease 2019 (COVID-19) spread globally and was declared a public health emergency of international concern on 30 January 2020 by the World Health Organization (1). Since then, the number of confirmed cases and deaths due to COVID-19 rapidly escalated, counting in millions, causing massive economic strain, and escalating healthcare and public health expenses (2, 3).

Response to the pandemic included a wide range of Public Health and Social Measures (PHSM). PHSM refer to a broad array of nonpharmaceutical interventions implemented by individuals, communities and governments to reduce the risk and scale of transmission of epidemic- and pandemic-prone infectious diseases. They range from individual measures such as hand washing, mask-wearing to social measures, such as physical distancing, modifying mass gathering events, school and business operations (4, 5). The World Health Organization has launched a multi-year initiative in 2021 to strengthen the global evidence base on PHSM and promote contextualized PHSM implementation (6).

In order to inform decisions by the public, health workforce and policy-makers, there is a need to synthesize the large volume of published work on COVID-19 (7). In the context of a high number of systematic reviews on this topic, an overview of systematic reviews becomes essential (8-10).

Existing overviews of systematic reviews on PHSMs for COVID-19 are either too narrow in the outcomes they address (e.g. focusing on symptoms and signs of COVID-19 in children and adolescents) (11) or too broad (e.g. analyzing systematic reviews published after the onset of the COVID-19 pandemic, up to March 24, 2020 and June 10, 2020, respectively) (10, 12). Furthermore, none of the to date existing overviews of systematic reviews explicitly addressed the unintended health, social and economic consequences of PHSMs (13).

### Review question

The proposed overview of systematic reviews is intended to synthesize the literature addressing the following:

- The effectiveness of single and combined PHSMs to reduce transmission of COVID-19
- The unintended desirable and undesirable impacts of the single and combined PHSMs addressing the COVID-19 pandemic on health outcomes and non-health outcomes including social, educational, and economic outcomes.

### Eligibility criteria

- **Studies designs:** We will include systematic reviews of primary studies of any design (experimental, observational and modelling studies) providing information on the effectiveness or unintended consequences of PHSMs in response to the COVID-19 pandemic. We will exclude abstracts, meetings proceedings, editorials and commentaries and?primary studies, protocols and literature reviews. We will also exclude reviews that did not include a risk of bias assessment of some kind as this is intended to help restrict the pool to reviews that were more likely to be well conducted and to contribute data that could be interpreted appropriately. For duplicate publications, we will select the most recent or complete version. Given the lack of consensus definition for a systematic review, Cochrane’s guidance for overviews of reviews recommends setting pre-established criteria for making decisions around inclusion. Thus, for this study, we define a systematic review as a publication where the following are provided: 1) an explicit search in at least two databases (at least one being electronic); 2) detailed description of the methods with explicit selection criteria; and 3) a summary of the included studies either in narrative or quantitative format (such as a meta-analysis). Both Cochrane and non-Cochrane systematic reviews will be considered eligible for inclusion, with or without meta-analysis, and regardless of language restriction and methodology of the included primary studies. Rapid reviews will be eligible for inclusion in this overview if they meet our pre-defined inclusion criteria noted above.
- **Population:** We will not restrict to any specific population as long as the study is in the context of the COVID-19 pandemic. We will exclude studies where we cannot isolate the data relevant to COVID-19.
- **Interventions**: The review will focus on PHSMs implemented in response to the COVID-19 pandemic. We will use the logic model developed by the WHO suggesting that PHSMs have two main objectives (14): to reduce the number of transmission-relevant contacts and to make contacts that take place safer. Based on a mapping of existing taxonomies, this would lead to 7 categories of PHSMs:
  - Surveillance (e.g. screening, testing, contact tracing)
  - Response (quarantine and isolation)
  - Services (school measures incl closures), business measures including closures, use of immunity/testing/vaccination certificates
  - Social interactions (e.g. physical distancing, stay at home orders, restrictions of mass gatherings)
  - Physical environment (e.g. ventilation, cleaning of objects and surfaces)
  - Individual protection (e.g. hand washing, respiratory etiquette, use of face masks)
  - Movement (e.g. travel restrictions including quarantine and testing and bans, restriction of domestic mobility)

We will include both single and multi-component interventions involving the eligible PHSM.

We will exclude reviews that only described PHSM interventions without reporting on any of the outcomes of interest as these cannot contribute to answering the research questions related to effectiveness and impact. We will also exclude reviews of studies that did not assess a specific PHSM or combination of PHSMs but rather looked at the extent of PHSMs overall, using the Oxford Scale. Additionally, we will exclude reviews that looked at indirect associations (for example, focusing on the impact of distance learning and digital media (which are a result of COVID-19 pandemic rather than PHSM) as well as reviews s. focusing on disinfectant interventions in clinical settings (versus public spaces)

### Outcome of interest

We will include systematic reviews reporting on any of the below outcomes (categorized according to the conceptual framework developed by the University of Munich and WHO^1^, derived from a review and mapping of existing PHSM taxonomies and literature (14):

#### Ttransmission-related outcomes

○ Epidemiologic outcomes, such as number of cases avoided, number of cases detected, positivity rate, change in outbreak pattern, transmission rates, morbidity and mortality rates from COVID-19

#### Health systems outcomes

○ These include healthcare utilization (e.g., hospitalization rates, number of cases requiring treatment in the intensive care unit (ICU), time until ICU capacity is reached), and health services availability (e.g., available intensive care units beds). Health systems outcomes

#### Non-COVID-19 related health outcomes

○ Non-COVID-19 related health outcomes include mental health, domestic violence, nutritional status/diet/BMI, substance use, sleeping patterns, physical activity, road accidents, and change in incidence and mortality of diseases other than COVID-19)

#### Unintended social and economic outcomes

○ These include homelessness/access to housing, social cohesion, educational attainment, days of schools missed, unemployment rates, economic productivity/growth, poverty, income levels, and financial support from government

#### Other outcomes (e.*g. environmental/ecological outcomes and human rights-related consequences)*

We will exclude reviews that include relevant studies that don’t report data in a way that these COVID study findings can be extracted separately. We will also exclude reviews focusing on the overall impact of COVID-19 pandemic on specific outcomes (e.g., physical inactivity) without specifying any particular intervention per se.

- **Setting**: We will include studies conducted in any country or setting (e.g. schools, businesses, points of entry, etc). We will not restrict eligibilty to any specific languages. We will exlcude reviews conducted in-vitro/laboratory setting (i.e. population of interest is not people or modeling of actual human settings)
- **Publication Venue**: We will exclude publications on preprint on servers such as medRxiv, bioRxiv, Litcovid and SSRN. This will help restrict the pool to higher quality reviews.

### Search methods for identification of reviews

The primary search source will be the Epistemonikos’ COVID-19 Living Overview of Evidence (LOVE) repository, a living repository that has been validated for use as a single source of COVID-19 evidence (15-17). The search will have no language restrictions. For the purpose of this overview of reviews, the search strategy will include specific keywords to represent the concept of ‘Public Health and Social Measures’ including the seven categories of PHSMs listed under ‘interventions’ (see eligibility criteria). The final list of terms was reviewed by a PHSM expert, and the final Boolean search strategy was reviewed by methods experts. The search strategy is available in Appendix 1.

Additionally, to detect potentially relevant systematic reviews in the COVID-10 LOVE repository, we will use a validated automatic classifier (a machine learning classifier for the records with an abstract) and a heuristic classifier for the records without an abstract (18). Finally, we will screen the reference lists of included reviews.

### Methodological assessment of included reviews

Two researchers will assess independently and in duplicate the quality of the included reviews using the AMSTAR tool 2 which enables more detailed assessment of systematic reviews that include randomized or non-randomized studies of healthcare interventions, or both (Shea et al., 2020). We will rate adherence to each item as follows: yes, partial yes, no, or not applicable (e.g. when a meta-analysis was not conducted). The overall confidence in the results of the review will be rated as “critically low”, “low”, “moderate” or “high”, according to the AMSTAR 2 guidance based on seven critical domains.

We will consider reviews that scored “low” or “very low” as having major limitations. We will also use the assessment of methodological limitation to choose between overlapping reviews.

Currently there is no guidance by the GRADE working group addressing the overview of reviews. We have proposed to abstract the GRADE certainty of evidence assessment by the review, if reported. If we identify at least one SR that is of moderate or high quality, we will choose the one with the highest quality and apply GRADE to its findings.

As indicated earlier, we will exclude all systematic reviews that don’t include a risk of bias assessment of some kind.

### Data extraction

Two researchers will extract data in duplicate and independently using standardized data abstraction form. They will resolve disagreements through consensus or with the help of a third reviewer if consensus could not be achieved. They will abstract data on the following variables from the included studies (to the extent that they have been provided by the included reviews):

- Type of synthesis document (e.g. standard versus rapid review)
- Basic information about systematic reviews (e.g. title; authors; year of publication; date last assessed as up-to-date; number of included studies; objectives; specific research question(s) (if explicitly provided by the authors), eligibility criteria, and study designs).
- Information on primary studies included in the systematic reviews: the range of primary study designs that the included reviews considered and identified in the review
- Context (countries/region, setting (e.g. healthcare settings, education facilities, etc.), phase of outbreak (if reported a priori in the methodology section), and population of interest at review level
- Interventions and comparators at review level (these will subsequently be categorized using the WHO logic model described earlier)
- Reported outcomes (as specified in Methods section of the systematic reviews).
- Key findings (including narratively reported study-level data and/or meta-analyzed data)
- Risk of bias assessment of included studies, if reported
- GRADE certainty of evidence assessment, if reported
- Whether risk of bias and GRADE certainty assessments were taken into account in interpreting the findings of the review
- Additional information (e.g. overview author’s comments, conclusion, systematic review limitations, reported disclosures of interest, funding source).

We will pilot-test data abstraction form prior to data abstraction.

### Data synthesis

We will report results in tabular formats and narratively. We will summarize outcome data as they are reported in the included systematic reviews, for both narratively reported study-level data, as well as meta-analyzed data.

For each outcome category (as specified by the WHO logic model), we will assess whether the intervention had a “desirable effect, little or no effect, an uncertain effect (very low□certainty evidence), no included studies, an undesirable effect, not reported (i.e. not specified as a type of outcome that was considered by the review authors), or not relevant (i.e. no plausible mechanism by which the type of intervention could affect the type of outcome)”. We will present the result in an intervention□outcome matrix.

### Managing overlapping systematic reviews

We will not exclude overlapping systematic reviews because, according to Cochrane’s guidance, it may be appropriate to include all relevant reviews’ results if the purpose of the overview is to present and describe the current body of evidence on a topic (9).

### Analysis of subgroups or subsets

None planned

## Data Availability

All data produced in the present work are contained in the manuscript

## Footnotes

1 The PHSM conceptual framework has since undergone extensive stakeholder consultation and review and is expected to be published on the WH PHSM website by the end of 2023.

## Notes

### Competing Interest Statement

The authors have declared no competing interest.

### Funding Statement

This review is funded by the NIPH

## References

1. WHO. WHO Emergencies Coronavirus Emergency Committee Second Meeting. 2020.

2. Brodeur A, Gray D, Islam A, Bhuiyan S. A literature review of the economics of COVID-19. Journal of Economic Surveys. 2021;35(4):1007–44.

3. Tandon PN. COVID-19: Impact on health of people & wealth of nations. The Indian journal of medical research. 2020;151(2-3):121.

4. WHO. Preparedness and Resilience for Emerging Threats 2023.

5. WHO. Considerations for implementing and adjusting public health and social measures in the context of COVID-19. 2023.

6. WHO. Measuring the effectiveness and impact of public health and social measures. [Available from: https://www.who.int/activities/measuring-the-effectiveness-and-impact-of-public-health-and-social-measures.

7. El-Jardali F, Bou-Karroum L, Fadlallah R. Amplifying the role of knowledge translation platforms in the COVID-19 pandemic response. Health Research Policy and Systems. 2020;18:1–7.

8. Bastian H, Glasziou P, Chalmers I. Seventy-five trials and eleven systematic reviews a day: how will we ever keep up? PLoS Med. 2010;7(9):e1000326.

9. Pollock M, Fernandes RM, Becker LA, Pieper D, Hartling L. Chapter V: overviews of reviews. Cochrane handbook for systematic reviews of interventions version. 2020;6.

10. Borges do Nascimento IJ, O’Mathúna DP, von Groote TC, Abdulazeem HM, Weerasekara I, Marusic A, et al. Coronavirus disease (COVID-19) pandemic: an overview of systematic reviews. BMC Infect Dis. 2021;21(1):525.

11. Viner RM, Ward JL, Hudson LD, Ashe M, Patel SV, Hargreaves D, et al. Systematic review of reviews of symptoms and signs of COVID-19 in children and adolescents. Archives of disease in childhood. 2021;106(8):802–7.

12. Hatmi ZN. A systematic review of systematic reviews on the COVID-19 pandemic. SN Comprehensive Clinical Medicine. 2021;3(2):419–36.

13. Regmi K, Lwin CM. Factors associated with the implementation of non-pharmaceutical interventions for reducing coronavirus disease 2019 (COVID-19): A systematic review. International journal of environmental research and public health. 2021;18(8):4274.

14. Rehfuess EA, Movsisyan A, Pfadenhauer LM, Burns J, Ludolph R, Michie S, et al. Public health and social measures during health emergencies such as the COVID-19 pandemic: An initial framework to conceptualize and classify measures. Influenza and Other Respiratory Viruses. 2023;17(3):e13110.

15. Verdugo-Paiva F, Vergara C, Avila C, Castro-Guevara JA, Cid J, Contreras V, et al. COVID-19 Living Overview of Evidence repository is highly comprehensive and can be used as a single source for COVID-19 studies. J Clin Epidemiol. 2022;149:195–202.

16. Pierre O, Riveros C, Charpy S, Boutron I. Secondary electronic sources demonstrated very good sensitivity for identifying studies evaluating interventions for COVID-19. J Clin Epidemiol. 2022;141:46–53.

17. Butcher R, Sampson M, Couban RJ, Malin JE, Loree S, Brody S. The currency and completeness of specialized databases of COVID-19 publications. J Clin Epidemiol. 2022;147:52–9.

18. Rada G, Perez D, Araya-Quintanilla F, Avila C, Bravo-Soto G, Bravo-Jeria R, et al. Epistemonikos: a comprehensive database of systematic reviews for health decision-making. BMC Med Res Methodol. 2020;20(1):286.

